# A Valid and Reliable Virtual Reality Tool for Assessment of Arm Nonuse Post-Stroke

**DOI:** 10.64898/2026.09.02.26361445

**Authors:** Rachana Gangwani, Shauna Zodrow, Alex DeAngelis, Maxim Karenbach, Shailesh Kantak, Laurel J. Buxbaum

**Author notes:** Corresponding Author: Rachana Gangwani, Ph.D. Address: 50 Township Line Road, Elkins Park, PA.

## Abstract

**Introduction:** Approximately 40-80% of individuals post-stroke fail to use their paretic arm despite sufficient capacity, a phenomenon known as *arm nonuse*. This behavior arises from an interaction between cognitive and motor factors, necessitating evaluation approaches that capture the impact of complex task demands on arm nonuse. This study evaluated the validity, reliability, and usability of a virtual reality-based arm nonuse (VR-NonUse) tool designed to measure arm choice during naturalistic reaching under varying cognitive and motor demands.

**Methods:** Fifty-one individuals with stroke and 23 controls completed the VR-NonUse assessment across three conditions varying in cognitive and motor demand. Arm choice was assessed on each trial. Usability and tolerability were evaluated. Concurrent validity was examined by assessing associations between paretic arm choice on the VR-NonUse tool and established nonuse assessments. Discriminant validity was evaluated by comparing these associations with a motor impairment measure. Construct validity was examined by determining the effects of cognitive and motor demand on paretic arm choice. Internal consistency reliability was assessed using a split-half reliability analysis with Spearman-Brown correction.

**Results:** The VR-NonUse tool was usable, well-tolerated, and demonstrated high internal-consistency reliability. Paretic arm choice on the VR-NonUse tool was associated with established nonuse measures, and more strongly related to nonuse than to motor impairment, supporting concurrent and discriminant validity. Higher cognitive demand reduced paretic arm choice, supporting construct validity. A brief cognitively demanding condition yielded a robust index of nonuse.

**Discussion:** The VR-NonUse tool is a reliable, valid, and practical assessment of arm nonuse in chronic stroke.

## Introduction

Approximately 40-80% of individuals after stroke exhibit *arm nonuse*, a phenomenon characterized by reduced use of the paretic upper extremity in daily life despite residual functional capacity.^1–3^ This dissociation between motor capacity and real-world arm use persists even following interventions that improve upper-extremity function. For example, high-dose task-specific interventions can improve motor capacity on standardized assessments, yet these gains do not necessarily translate to increased paretic arm use in real-world settings.^4,5^ The distinction between capacity and use has important implications for post-stroke outcomes, as post-stroke participation and quality of life are more strongly associated with real-world arm use than with motor impairment or capacity alone.^6^ Collectively, these findings underscore the need for approaches that capture arm nonuse under conditions that approximate everyday demands.

Growing evidence suggests that arm nonuse reflects factors beyond motor impairment alone. Individuals with similar impairments often exhibit different patterns of paretic arm use,^7^ and relatively preserved motor function does not guarantee greater arm use.^8–10^ Rather, arm use may reflect the integration of multiple factors, including sensorimotor processing, cognitive demands, attentional/arousal states, affective influences, and habit.^11–14^ Contemporary models conceptualize arm use as an implicit action-selection process in which the sensorimotor system weighs the expected physical and cognitive costs and benefits of using one arm versus the other.^13,14^ Consistent with this framework, our prior research using a virtual reality (VR) reaching task showed that increasing cognitive demands exacerbated paretic arm nonuse even when motor demands remained constant.^15,16^

Current methods for assessing arm nonuse remain limited. Observational measures such as the Actual Amount of Use Test (AAUT) quantify arm use across predefined tasks.^17,18^ While valuable, the AAUT requires specialized materials, examiner training, and covert video recording, limiting its clinical feasibility. Wearable accelerometers provide objective measures of real-world arm movement frequency^19,20^ but offer little insight into the context of arm use and involve substantial cost and data processing. Other approaches assess arm use behavior during structured tasks. For example, the Bilateral Arm Reaching Test used a two-dimensional reaching task to quantify arm use,^21^ whereas the Rehabilitation Gaming System utilized goal-directed reaching movements in a virtual environment to investigate acquired nonuse.^22^ Although these approaches have advanced the assessment and understanding of arm nonuse, they do not systematically evaluate the effects of varying cognitive and motor task demands, as encountered in daily real-world situations, on spontaneous arm choice. Thus, there remains a need for clinically feasible assessment tools that quantify arm choice under different task demands.^23^Virtual reality (VR) provides a promising solution to addressing this need by enabling the assessment of spontaneous arm choice during standardized, goal-directed tasks while systematically manipulating task demands. By controlling task instructions, stimulus presentation, and response requirements, VR allows the effects of specific task characteristics on arm choice to be examined under consistent conditions. This approach may provide insight into the factors influencing spontaneous arm selection beyond measures of overall arm-use frequency. Building on our prior work demonstrating the influence of cognitive demands on paretic arm choice during goal-directed reaching,^15^ we developed the Virtual Reality Nonuse tool (VR-NonUse), a modified version of our previously described VR-based assessment. The VR-NonUse tool assesses arm choice on each trial by recording whether participants select their paretic or nonparetic arm to complete the task. The proportion of trials in which participants select the paretic arm are then used to quantify paretic arm use. The revised VR-NonUse tool was designed to improve ease of administration and facilitate future clinical implementation while extending the assessment to incorporate systematic manipulation of both cognitive and motor demands on arm choice. Cognitive demands were manipulated by requiring selection of target objects from a visual array containing either different-category distractors (lower cognitive demand) or same-category distractors (higher cognitive demand). Motor demands were manipulated by requiring responses comprising either “poking” or grasping target objects.

The aim of this study was to establish the psychometric properties of a research version of the VR-NonUse tool. Specifically, we evaluated its concurrent, discriminant, and construct validity, as well as its internal consistency reliability, usability, and tolerability. The study was informed by a framework describing distinct phases of clinical tool development, in which Scale Development is characterized as Phase 2.^24^ We hypothesized that the VR-NonUse tool would demonstrate evidence of validity and reliability for assessing arm nonuse following stroke. Specifically, we expected that (a) the proportion of paretic arm choice on the VR-NonUse tool would be associated with established measures of arm nonuse, (b) these associations would be stronger than with motor impairment and (c) paretic arm choice would vary with task demands. We further hypothesized that the tool would demonstrate acceptable internal consistency reliability, usability, and tolerability. Although the VR-NonUse tool also captures movement performance metrics (e.g., kinematics), this study focused on validating its assessment of arm nonuse.

## Methods

### Participants

Fifty-one right-handed individuals with stroke (29 left cerebrovascular accident (LCVA); 22 right cerebrovascular accident (RCVA); 18 females, mean age=60.4±12.1 years; time post-stroke=67.6±67.9 months) and 23 age-matched right-handed neurotypical individuals (12 females, mean age=64.4±12.6 years) were recruited from the Neurocognitive Rehabilitation Research Registry at Moss Rehabilitation Research Institute.^25^

Inclusion criteria were age 18-90 years and the ability to provide informed consent. Individuals with stroke were required to have a single unilateral stroke, be ≥6 months post-stroke, have sufficient auditory and visual acuity (with or without corrective lenses) to identify computer-based stimuli, and demonstrate the ability to lift the paretic arm to at least chest height (Upper Extremity Fugl Meyer [UEFM] score ≥30).

Exclusion criteria included significant neurological or medical conditions (other than stroke in the stroke group), poorly controlled psychiatric disorders, current alcohol or drug abuse, and contraindication to virtual reality (e.g., cybersickness). Susceptibility to cybersickness was assessed through self-report rather than a standardized questionnaire. Specifically, potential participants were asked about their previous VR experiences and whether they had experienced discomfort or other adverse effects during VR exposure. Individuals with stroke were excluded if they exhibited severe language comprehension impairment (Western Aphasia Battery auditory verbal comprehension score <4), or upper-extremity injury/pain that would interfere with reaching movements. Individuals with stroke exhibiting mild-moderate aphasia, spasticity, spatial neglect, visual extinction, or somatosensory and proprioceptive deficits were not excluded to represent the range of impairments commonly observed post-stroke. The influence of these factors on arm nonuse is being investigated in a companion study. Control participants were excluded for cognitive impairment (Montreal Cognitive Assessment score <16).^26^ Participant demographics and behavioral assessment scores are shown in Table 1.

**Table 1:** Participant demographics and behavioral assessment scores.

| ID | Edu.<br>(yrs) | Sex | MPO | UEFM | AAUT<br>Nonuse<br>Index | Use<br>Ratio | Basic<br>Grasp | Complex<br>Grasp | Complex<br>Point |
|---|---|---|---|---|---|---|---|---|---|
| RCVA1 | 17 | M | 65 | 59 | 0.58 | 0.72 | 1.6 | 11.1 | 11.5 |
| RCVA2 | 12 | F | 41 | 62 | 0.07 | 1.25 | 50.3 | 52 | 0 |
| RCVA3 | 12 | M | 15 | 63 | 0.5 | 0.64 | 31.6 | 19 | 8.8 |
| RCVA4 | 14 | M | 188 | 61 | 0.84 | 0.98 | 43.1 | 58.2 | 83.9 |
| RCVA5 | 17 | M | 106 | 63 | 0.1 | 0.88 | 45.3 | 41.4 | 43.2 |
| RCVA6 | 18 | M | 18 | 52 | 0.66 | 0.35 | 35.1 | 16.8 | 35.3 |
| RCVA7 | 16 | M | 133 | 48 | 0.64 | 0.43 | 64.7 | 64.7 | 62.2 |
| RCVA8 | 12 | M | 41 | 56 | 0.36 | 0.92 | 50.3 | 51.6 | 36.2 |
| RCVA9 | 9 | M | 19 | 65 | 0.21 | 0.79 | 37.2 | 37.8 | 33.5 |
| RCVA10 | 16 | M | 29 | 55 | 0.71 | 0.65 | 36.2 | 28.2 | 30.5 |
| RCVA11 | 16 | M | 21 | 30 | 1 | 0.36 | 26 | 19.2 | 53.2 |
| RCVA12 | 16 | M | 52 | 55 | 0.73 | 0.44 | 2.6 | 4.8 | 6.3 |
| RCVA13 | 18 | F | 7 | 60 | 0.43 | 0.66 | 42.6 | 39.9 | 45 |
| RCVA14 | 12 | M | 69 | 58 | 0.36 | 0.64 | 32.5 | 33 | 35.1 |
| RCVA15 | 12 | F | 56 | 56 | 0.86 | 0.6 | 47.6 | 30.9 | 45 |
| RCVA16 | 12 | M | 28 | 38 | 1 | 0.57 | 8.6 | 5.4 | 18.3 |
| RCVA17 | 12 | M | 17 | 35 | 1 | 0.53 | 48.4 | 47 | 48.7 |
| RCVA18 | 14 | M | 61 | 55 | 0.43 | 0.56 | 49.5 | 45.9 | 39.2 |
| RCVA19 | 12 | M | 23 | 55 | 0.54 | 0.72 | 23.4 | 36.5 | 28.3 |
| RCVA20 | 14 | M | 175 | 51 | 0.36 | 0.62 | 45.8 | 26.3 | 24.1 |
| RCVA21 | 16 | F | 12 | 66 | 0.07 | 0.88 | 41.9 | 42.4 | 39.1 |
| RCVA22 | 14 | M | 9 | 43 | 0.5 | NA | 31.6 | 35.4 | 33.2 |
| Mean<br>(SD) | 14.1<br>(2.4) |  | 53.8<br>(52.1) | 53.9<br>(9.7) | 0.5<br>(0.2) | 0.67<br>(0.21) | 35.9<br>(16.2) | 34.8<br>(17.75) | 34.5<br>(19.1) |
| LCVA1 | 12 | F | 244 | 66 | 0.25 | 1.19 | 62 | 35.1 | 57.3 |
| LCVA2 | 18 | M | 77 | 58 | 0.28 | 0.55 | 18.8 | 2.4 | 0.6 |
| LCVA3 | 18 | F | 163 | 66 | 0.07 | 1.1 | 52.1 | 55.1 | 47.1 |
| LCVA4 | 12 | M | 190 | 66 | 0.3 | 1.09 | 56.5 | 48 | 43.3 |
| LCVA5 | 12 | M | 221 | 64 | 0.5 | 0.55 | 24.6 | 35.3 | 30.6 |
| LCVA6 | 14 | M | 185 | 60 | 0.35 | 0.6 | 24.3 | 4.8 | 23.8 |
| LCVA7 | 14 | M | 110 | 40 | 0.69 | 0.54 | 47.9 | 47.9 | 46.5 |
| LCVA8 | 16 | F | 33 | 66 | 0.14 | 1.24 | 65.6 | 64.2 | 69.4 |
| LCVA9 | 16 | F | 10 | 64 | 0.21 | 1.01 | 46.9 | 66.5 | 36 |
| LCVA10 | 13 | M | 57 | 65 | 0.41 | 1 | 50.3 | 47.4 | 46.1 |
| LCVA11 | 18 | M | 188 | 66 | 0.42 | 0.97 | 49.7 | 50 | 53.4 |
| LCVA12 | 16 | M | 61 | 40 | 0.53 | 0.53 | 35.5 | 43.1 | 35.7 |
| LCVA13 | 12 | M | 80 | 59 | 0.35 | 0.83 | 50.3 | 44.6 | 45.9 |
| LCVA14 | 13 | M | 36 | 58 | 0.08 | 0.98 | 53.1 | 56.3 | 56.8 |
| LCVA15 | 9 | M | 230 | 61 | 0 | NA | 59.4 | 54.5 | 57.7 |
| LCVA16 | 12 | M | 44 | 56 | 0.14 | 1.02 | 62.4 | 65.6 | 62.7 |
| LCVA17 | 16 | M | 21 | 57 | 0.57 | 0.74 | 50.3 | 49.5 | 46.4 |
| LCVA18 | 14 | F | 45 | 62 | 0.33 | 0.87 | 68.6 | 65.6 | 66 |
| LCVA19 | 12 | F | 15 | 66 | 0.07 | 1.08 | 49 | 50.3 | 50.6 |
| LCVA20 | 14 | F | 43 | 36 | 1 | 0.69 | 26.8 | 15.8 | 25 |
| LCVA21 | 18 | M | 10 | 54 | 0.77 | 0.55 | 51.6 | 51.1 | 53.7 |
| LCVA22 | 16 | F | 6 | 44 | 0.29 | 0.5 | 46.8 | 45.8 | 45.8 |
| LCVA23 | 18 | M | 8 | 46 | 0.64 | 0.68 | 57.3 | 50.6 | 51.6 |
| LCVA24 | 18 | F | 77 | 65 | 0.07 | 0.72 | 49.7 | 53.4 | 46 |
| LCVA25 | 20 | F | 9 | 61 | 0.14 | 1.05 | 52.4 | 51.9 | 51.6 |
| LCVA26 | 16 | F | 6 | 66 | 0.14 | 0.73 | 52.1 | 56.8 | 58.9 |
| LCVA27 | 14 | F | 22 | 61 | 0.57 | 0.77 | 64.9 | 48.1 | 54.1 |
| LCVA28 | 12 | F | 45 | 65 | 0.21 | 0.8 | 68.2 | 70.3 | 66.7 |
| LCVA29 | 20 | F | 30 | 56 | 0.25 | 0.7 | 44.8 | 61.8 | 50 |
| Mean<br>(SD) | 14.9<br>(2.8) |  | 78.1<br>(77.1) | 58.4<br>(8.8) | 0.3<br>(0.2) | 0.8<br>(0.2) | 49.7<br>(13) | 47.6<br>(17) | 46.6<br>(15.7) |
| CTL | 15.5<br>(2.5) | 12F,<br>11<br>M |  |  |  |  | 52.2<br>(12.4) | 49<br>(11.3) | 47.8<br>(9.2) |

Edu. (yrs): Years of education, M: Male; F: Female;MPO: Months post-stroke, UEFM: Upper Extremity Fugl Meyer, AAUT: Actual Amount of Use Test, RCVA: Right cerebrovascular accident, LCVA: Left cerebrovascular accident, CTL: Controls. NA: Data not available due to technical errors, SD: Standard deviation. Last three columns indicate proportion of paretic arm use calculated as the percentage of trials where the paretic arm was chosen in each of the three conditions.

### Procedure

#### Actual Amount of Use Test (AAUT)

The Actual Amount of Use Test (AAUT)^17^ required individuals with stroke to perform 14 upper-extremity tasks under two conditions. In the spontaneous condition, participants received no instructions regarding arm use whereas in the forced condition, they were instructed to use their paretic arm. Performance was video recorded throughout; participants were unaware of the recording during the spontaneous condition and were debriefed after the assessment. Tasks were scored dichotomously (0=no paretic arm use; 1=any paretic arm use), yielding a maximum score of 14 per condition. We measured the Nonuse Index,^18^ as the difference in arm use between the forced and spontaneous conditions, normalized to arm use in the forced condition [Nonuse Index = (arm use in forced condition – arm use in spontaneous condition) / (arm use in forced condition)]. Scores ranged from 0 (no nonuse) to 1 (complete paretic arm nonuse).

#### Actigraph Use Ratio

Participants wore bilateral wrist-mounted accelerometers (ActiGraph GT9X Link, Pensacola, FL, USA) during waking hours for five consecutive days to quantify real-world arm use. Triaxial acceleration (0.001664g/count) was sampled at 30Hz and aggregated into 1-second epochs using ActiLife™ 6 software. Data were processed in MATLAB (MathWorks Inc., Natick, MA, USA) using custom-written algorithms. For each arm, activity duration was calculated as the number of 1-second epochs with a nonzero activity value (i.e., activity > 0). The Use Ratio,^27^ was calculated as paretic arm activity duration divided by the non-paretic arm activity duration. A Use Ratio of 1 indicates equal activity duration of the paretic and non-paretic arms, whereas values below or above 1 indicate relatively less or greater paretic arm use, respectively; a value of 0 indicates complete paretic arm nonuse.

#### Virtual Reality Nonuse (VR-NonUse) Tool

##### Target familiarization

Prior to the assessment, participants were familiarized with the target objects. Two-dimensional black-and-white and colored three-dimensional images of objects were drawn from three semantic categories (household items, animals, and produce (fruits/vegetables); 16 objects/category), and were presented individually on a computer, along with their written and spoken names.

##### Apparatus and display

Participants wore a Meta Quest 2 head-mounted display with integrated headphones and hand tracking which presented an immersive, three-dimensional virtual environment. They were seated with their hands in start positions on an adjustable table at lap height. Participants were instructed to reach toward previously familiarized objects presented on three shelves of a virtual shelving unit located ∼60cm in front of the lap tray (adjusted for participants’ arm length), subtended ∼66.7° of visual angle, with shelves positioned 6, 21, and 36cm above the lap tray. On each trial, 12 three-dimensional appearing objects appeared in a 4-column x 3-row grid of the shelving unit. Two avatar hands were presented from a first-person perspective (See Figure 1A) and synchronized with participants’ real-time hand movements to provide visual feedback of hand position within the VR environment.

**Figure 1.**
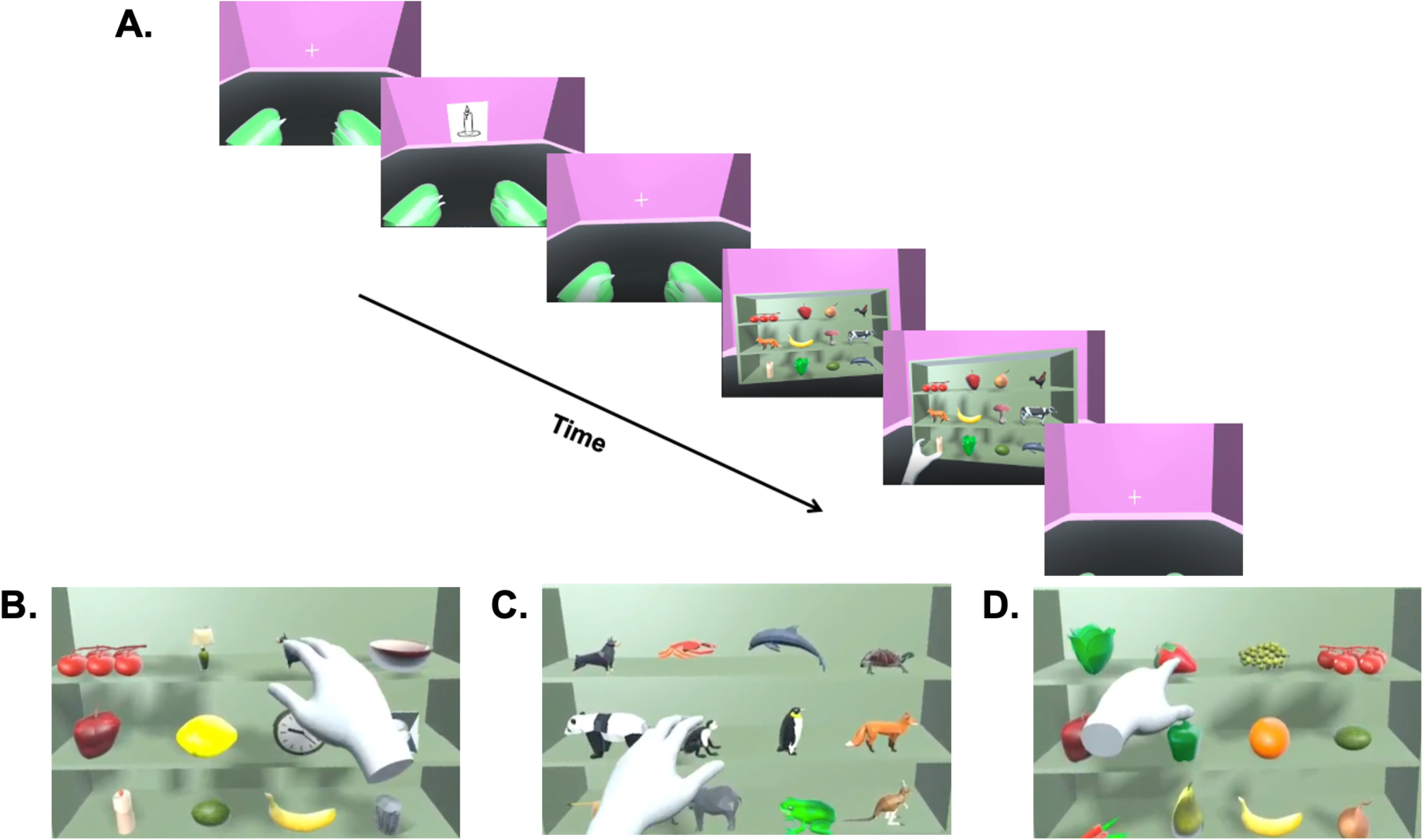
Screenshots of the Virtual Reality Nonuse (VR-NonUse) tool. A: Schematic illustration of the sequence of events within a single trial. B–D: Representative examples of the Basic Grasp (B), Complex Grasp (C), and Complex Point (D) conditions.

##### Practice trials

Participants were instructed to touch the four corners of the virtual shelf and to reach to a ball presented at each target location to confirm they could reach all possible locations with both arms. Participants completed 10 practice trials involving reaching with either arm to point or grasp target objects among semantically related distractors.

##### Trial events

Each trial (Figure 1A) began after participants signaled their readiness by placing their hands in the start positions, denoted by colored orbs that appeared ∼8 inches to the left and right of midline. A central fixation cross then appeared for 1000ms, followed by the spoken target word and a 2-dimensional black-and-white line drawing of the target object presented for 2000ms. A second central fixation cross then appeared for a variable interval (1000-2000ms) followed by presentation of the shelving unit containing 12 objects. Participants had 7000ms to locate and touch or grasp the target object.^15,28^ Before each condition, participants were instructed to freely choose either arm to reach and poke or grasp to lift the target object and to complete the task as quickly and accurately as possible. Trials in which participants reached for or grasped an incorrect target, or otherwise failed to complete the task accurately, were excluded from the analyses. Although participants could see the movements of their avatar hands, no feedback indicating trial success or accuracy, or regarding arm choice, was provided during the assessment.

##### VR-NonUse Tool conditions

Participants were tested under three conditions. In the Basic Grasp condition (Figure 1B), participants reached to grasp and lift a target object from different semantic categories than the distractors (e.g., target=animal, distractors=household objects). In the Complex Grasp condition (Figure 1C), participants reached to grasp and lift a target object from the same semantic category (e.g., target=animal, distractors=other animals), increasing search difficulty through visual and semantic competition.^29,30^ Lifting of the target object could be accomplished with any type of grip (e.g., precision or power), but loss of contact between the object and the shelf surface was required for the trial to be considered accurate. In the Complex Point condition (Figure 1D), participants reached to poke the target object, presented among distractors from the same semantic category, using any finger or combination of fingers.

##### Experimental design

Each of the 12 array positions served as the target location on 8 trials in each condition, yielding 24 trials/column, and 96 trials/block. Participants performed 2 blocks per condition (6 blocks total; 576 trials). Distractor objects were randomly allocated to array positions and each object appeared twice as a target in each condition at different positions. Condition order was counterbalanced using a Latin Square design.

##### Outcome measures

The primary outcome measure was arm choice on each trial, defined as the spontaneous selection of the paretic arm to complete the task. For each condition, paretic arm use on the VR-NonUse tool was calculated as the number of valid trials performed with the paretic arm in each condition divided by the total observations in that condition, multiplied by 100. As noted, the VR-NonUse tool also enables analyses of reaction time, movement time, and reach kinematics, which will be reported in a subsequent manuscript.

##### Test sessions

Individuals with stroke completed 5-6 visits over approximately 6 weeks. At Visit 1, participants completed the AAUT, along with measures of stroke severity (NIH Stroke Scale) and a measure of lateralized and non-lateralized visual attention (Virtual Reality Lateralized Attention Test-2; VRLAT-2), that shows evidence of moderate discriminant and concurrent validity.^31,32^ Visit 2 included sensorimotor assessments. During Visits 3-5/6, all participants completed the 6-block VR-NonUse assessment (two blocks x three conditions), performing 2 or 3 blocks per session according to their assigned randomization sequence. Thus, the full VR-NonUse assessment was completed over two or three sessions. Each block required approximately 12-15 minutes to complete. Each session lasted approximately 60-90 minutes with rest breaks provided between blocks and as needed to accommodate participant fatigue. At Visit 3, participants received accelerometers for approximately one week. Control participants completed 2-3 sessions that included the VRLAT and the VR-NonUse assessment.

#### Assessments of usability and tolerability

Subjective measures of usability and simulator-related discomfort (i.e., cybersickness) were administered during the final session. Perceived usability of the VR-NonUse Tool was assessed using the 10-item System Usability Scale (SUS),^33^ with 5 response options (strongly disagree to strongly agree). Responses were scored from 0 to 4 and multiplied by 2.5 resulting in a composite score of 0 to 100, with higher scores indicating greater usability. We also administered the Simulator Sickness Questionnaire (SSQ),^34^ a 16-item measure of discomfort, fatigue, nausea, and other cybersickness-related symptoms whose severity was rated on a 4-point scale (0=none to 3=severe). Total scores <5 indicate minimal symptoms, whereas scores >20 reflect poor tolerability.

### Statistical analysis

Descriptive statistics of SUS and SSQ scores were calculated to evaluate usability and tolerability.

Concurrent validity was assessed by examining correlations between the proportion of paretic arm choices in each VR-NonUse condition and established measures of arm nonuse and real-world arm use (AAUT Nonuse Index and the ActiGraph Use Ratio respectively). To account for multiple comparisons, p-values were separately adjusted using the Benjamini-Hochberg false discovery rate (FDR) procedure.^35^

Discriminant validity was evaluated by comparing the associations of the proportion of paretic arm choices during each VR-NonUse condition with established measures of arm nonuse and real-world arm use versus its association with motor impairment (UEFM). Differences in correlation strength were tested using Steiger’s test, as correlations were estimated within the same sample and shared common variance.^36^ For these comparisons, the signed correlation coefficients were entered into the Steiger tests with the scoring direction of each comparator measure retained with p-values adjusted using the Benjamini-Hochberg FDR procedure.^35^ Correlation coefficients were interpreted based on their absolute magnitude, with values <0.40 considered weak, 0.40-0.70 moderate, 0.70-0.90 strong, and 0.90-1.00 very strong.^37^

Construct validity was examined using linear mixed-effects models assessing the effect of motor and cognitive demand on paretic (stroke group) and non-dominant (control group) arm choice counted on a trial level. Models included fixed effects of Condition (Basic Grasp, Complex Grasp, Complex Point), Group (RCVA, LCVA, and controls), and the Group × Condition interaction, with Subject included as a random effect. Stroke participants were labeled by lesioned hemisphere based on evidence indicating hemispheric differences in paretic arm use.^15,38^ The assumption of homogeneity of variance was evaluated by examining model residuals for evidence of systematic changes in residual variance across fitted values and experimental groups/conditions. The residual diagnostics did not indicate substantial heteroscedasticity. Post hoc comparisons following the mixed-effects analyses were performed using estimated marginal means with Tukey adjustment for multiple comparisons. These included comparisons between conditions and between groups.

Internal consistency reliability of the VR-NonUse tool was evaluated separately for each condition. Using a split-half analysis, valid trials were divided into odd- and even-numbered trials based on their original sequence, and the proportion of paretic/non-dominant arm choice was calculated separately for each half for each participant. Spearman correlations were calculated between odd and even trials, and Spearman–Brown correction was applied to estimate the reliability of scores based on the full set of trials within each condition.^39–41^

To support development of a clinically feasible short version of the tool, an additional mixed-effects model examined the effect of block repetition. This model included fixed effects of Condition, Block within Condition (Blocks 1 and 2), and the Condition × Block interaction, with Subject specified as a random effect.

To develop a preliminary approach for identifying individuals with stroke exhibiting arm nonuse, receiver operating characteristic (ROC) analysis was performed to evaluate how well paretic arm choice during the Complex Grasp condition distinguished individuals who showed nonuse on the AAUT from those who did not show nonuse on the AAUT, based on the AAUT Nonuse Index. Although there is no “gold standard” for determining nonuse, we used the published AAUT Nonuse Index as the reference measure, with values >0.5 classified as nonuse and values ≤0.5 classified as no nonuse, based on a threshold employed in previous work.^9^ The proportion of paretic arm choice in the Complex Grasp condition was entered as the continuous predictor, with AAUT-defined nonuse status as the binary outcome. Discriminative ability was quantified using the area under the curve (AUC). An exploratory threshold for paretic arm choice was selected by maximizing sensitivity and specificity (Youden’s index; sensitivity + specificity – 1).^42^ Because the threshold was derived from the present sample, it was considered preliminary and requires independent validation.

The present study was conducted as part of a broader, multi-aim study, and no formal a priori power analysis was conducted specifically for these validation analyses. Sample size was determined by balancing feasibility considerations with the goal of completing this phase of tool development and making the VR-NonUse tool available for further evaluation in a timely manner. Post hoc sensitivity analyses were conducted to characterize the statistical sensitivity of the study design. For the correlation-based validity analyses, an approximate sensitivity analysis was conducted using the sample of 51 participants with stroke, a two-sided α=.05, and 80% power.^43^ For the mixed-effects construct-validity analysis, simulation-based sensitivity analyses were conducted using the fitted linear mixed-effects model as the data-generating model.^44,45^ The model included fixed effects of Group, Condition, and the Group × Condition interaction and a random intercept for participant. Fixed-effect estimates and variance components from the model fitted to the full observed sample were held constant across simulations. We varied the number of simulated participants (*N*=30, 40, 50, 60, and 74) to examine how power changed with sample size, while approximately preserving the observed group composition and repeated-trial structure. For each sample size, 1,000 datasets were simulated and analyzed using the same linear mixed-effects model as the primary analysis. Estimated marginal condition means and Tukey-adjusted pairwise comparisons were then used to evaluate the cognitive-demand contrast (Basic Grasp vs. Complex Grasp) and motor-demand contrast (Complex Grasp vs. Complex Point). Power was estimated as the proportion of simulated datasets in which the respective comparison yielded *p*<.05. Because these analyses were conducted post hoc and incorporated effect estimates and variance components obtained from the observed data, they were interpreted as descriptive assessments of sensitivity to effects of the magnitude observed in the present sample rather than as retrospective justification of the original sample size.

## Results

### Usability and Tolerability

All included participants completed the VR-NonUse assessment across the scheduled study visits. SUS scores were high in both groups (controls: 78.9±10; stroke: 80.2±14.2), indicating good-to-excellent usability.^33^ Average SSQ scores were low for both controls (18.3±2.1) and the stroke group (19.1±3.4), indicating low simulator sickness^34^ and suggested that the VR-NonUse assessment was well-tolerated by both groups.

### Concurrent validity

Paretic arm choice on the VR-NonUse tool was negatively correlated with the AAUT NonUse Index in the Basic Grasp (ρ=−0.40, p=0.003, q=0.004) and Complex Grasp (ρ=−0.46, p=0.0006, q=0.001) conditions but not in the Complex Point condition (ρ=−0.20, p=0.15, q=0.15) (Figure 2A). Paretic arm choice on the VR-NonUse tool was positively correlated with the Actigraph Use Ratio across all conditions (Basic Grasp: ρ=0.53, p<0.0001, q=<0.00015; Complex Grasp: ρ=0.52, p<0.0001, q=<0.00015; Complex Point: ρ=0.32, p=0.02, q=0.02) (Figure 2B). Together, these findings indicate that participants who demonstrated less paretic arm use during the VR-NonUse assessment generally exhibited greater nonuse on the AAUT and lower paretic arm use in daily life, with the most consistent associations observed in the Basic Grasp and Complex Grasp conditions.

**Figure 2.**
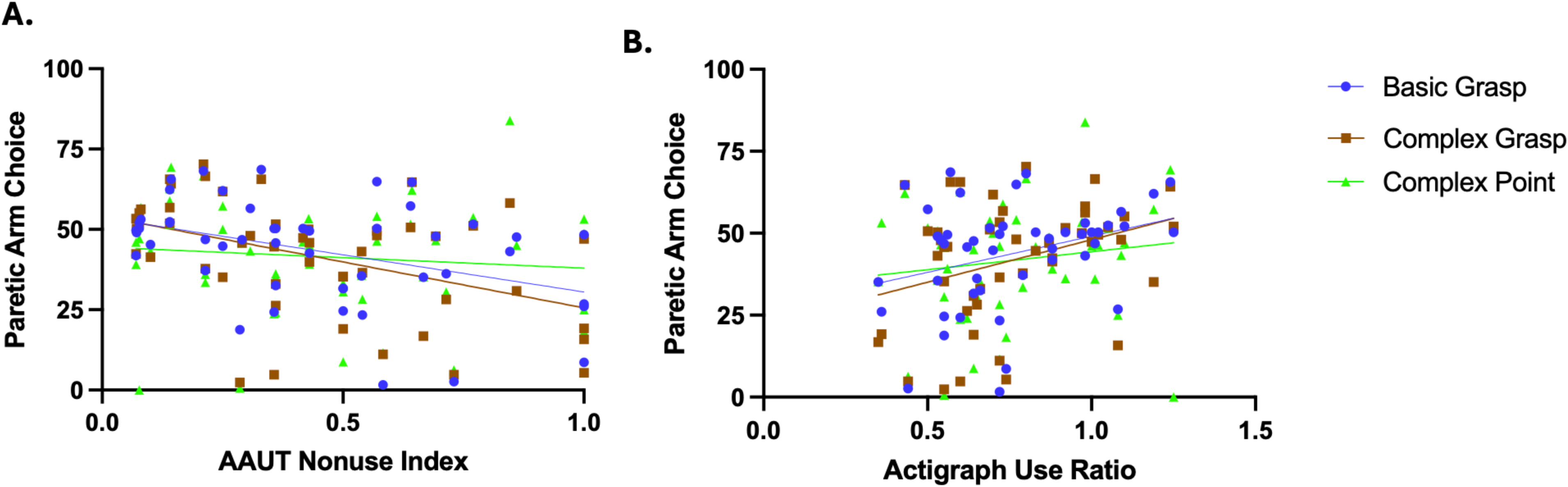
Relationship between proportion of paretic arm choice in each VR-NonUse assessment condition with (A) Actual Amount of Use Test (AAUT) Nonuse Index and (B) Actigraph Use Ratio.

### Discriminant validity

Paretic arm choice on the VR-NonUse tool was numerically more strongly associated with the AAUT Nonuse Index and Actigraph Use Ratio than with UEFM (Supplemental Material Table 1). Steiger’s tests indicated that associations between the VR-NonUse tool and AAUT Nonuse Index were significantly stronger than associations with the UEFM in the Basic Grasp (z=3.11, p=0.002) and Complex Grasp (z=3.38, p<0.001) conditions, but not in the Complex Point condition (z=1.67, p=0.09). Similarly, Steiger’s tests confirmed that correlations between the VR-NonUse tool and Actigraph Use Ratio were significantly stronger than those with the UEFM in the Basic Grasp (*z*=1.96, *p*=0.05) and Complex Grasp (*z*=1.94, *p*=0.05) conditions but not in the Complex Point condition (*z*=0.88, *p*=0.38). These findings indicate that paretic arm choice measured during the Basic Grasp and Complex Grasp conditions was significantly more closely associated with other established measures of arm nonuse and real-world arm use (AAUT Nonuse Index and Actigraph Use Ratio respectively) than with upper-extremity motor impairment, suggesting that paretic arm choice in the VR-NonUse tool captures aspects of arm use behavior not fully reflected by motor impairment scores.

### Construct validity

Our model assessing the effects of Group and Condition on paretic (stroke group) or non-dominant (control) arm choice on each trial revealed a significant main effect of Group (*F*_(2,41159)_=334.37, *p*<0.0001) and Condition (*F*_(2,41167)_=17.24, *p*<0.0001). The Group × Condition interaction was not significant (*F*_(2,41167)_=1.15, *p*=0.32). Post hoc analyses demonstrated that the proportion of paretic/non-dominant arm choice was lower in the Complex Grasp than the Basic Grasp condition (t_(41167)_=3.96, *p*=0.0002), indicating reduced paretic arm use as cognitive demand increased. Arm choice did not differ between the Complex Grasp and Complex Point conditions (t_(41167)_=1.70, *p*=0.18), indicating no effect of the motor-demand manipulation (Figure 3). Compared with controls’ non-dominant arm, paretic arm choice was lower in both the LCVA (t_(41158)_=8.93, *p*<0.0001) and RCVA groups (t_(41158)_=16.00, *p*<0.0001). Paretic arm choice was significantly lower in the RCVA than the LCVA group (t_(41158)_=25.26, *p*<0.0001). Overall, participants with stroke used their paretic arm less frequently than controls used their non-dominant arm, with the lowest proportion of paretic arm use observed in the RCVA group.

**Figure 3.**
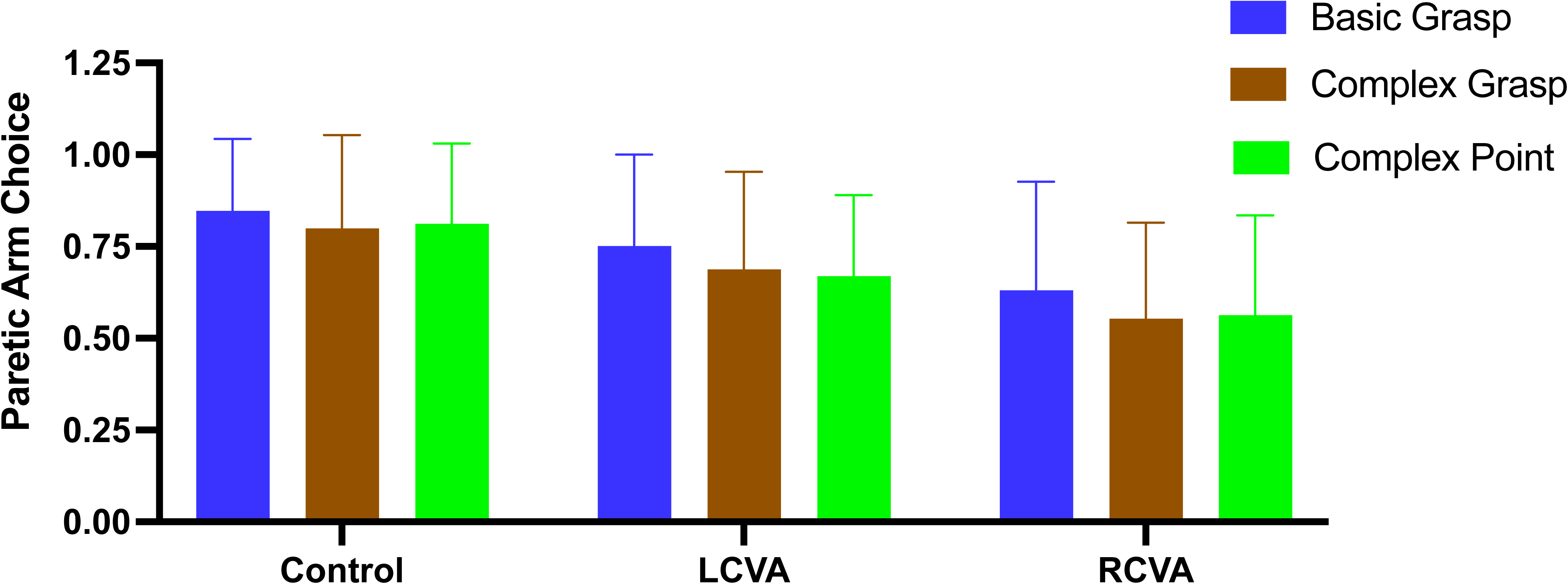
Proportion of paretic/non-dominant arm choice across the VR-NonUse assessment conditions in individuals with left and right hemispheric stroke (LCVA and RCVA) and controls.

### Internal Consistency Reliability

Arm choice proportions demonstrated high internal consistency reliability across all three VR-NonUse conditions. The proportion of paretic/non-dominant arm choices calculated from odd- and even-numbered trials were strongly correlated in the Basic Grasp (*r*=0.88), Complex Grasp (*r*=0.94), and Complex Point (*r*=0.92) conditions. The corresponding Spearman–Brown corrected reliability coefficients were 0.93, 0.97, and 0.96, respectively. This indicates that the measure has internal stability.

### Item Reduction Analysis

An important stage in clinical test development is item reduction analysis (Phase 2, Step 5).^24^ Toward this end, we examined differences between first versus second block of each condition using a linear mixed-effects model with fixed effects of Condition, Block, and their interaction. The model revealed a significant main effect of Condition (*F*_(2,41171)_=16.31, *p*<0.0001) and a significant Condition × Block interaction (*F*_(2,41171)_=3.23, *p*=0.03), but no significant main effect of Block (*F*_(1,41171)_=2.7, *p*=0.09) on paretic arm choice. Post hoc analyses of the interaction showed no significant differences between the first and second blocks of the Basic Grasp (*t_(41171)_*=2.44, *p*=0.14), Complex Grasp (*t_(41171)_*=1.53, *p*=0.64) or Complex Point (*t_(41171)_*=−1.06, *p*=0.89) conditions. The significant interaction therefore reflected a numerically larger, but non-significant, difference between the first and second blocks in the Basic Grasp condition compared with the other conditions (Supplemental Material Figure 1). The absence of block effects supports development of a shortened version with one block per condition.

To explore a preliminary criterion for identifying arm nonuse on the VR-NonUse tool, ROC analysis was performed across all 51 participants with stroke, of whom 18 showed nonuse on the AAUT and 33 did not show nonuse on the AAUT, based on the AAUT Nonuse Index. ROC analysis demonstrated moderate discrimination between these groups based on spontaneous paretic arm choice during the first Complex Grasp block (AUC=0.664). The threshold maximizing Youden’s index was 30.9% paretic arm choice, yielding an overall classification accuracy of 72.5%, sensitivity of 44.4%, specificity of 87.9%, and Youden’s index of 0.323. At this threshold, 8 out of 18 participants (44.4%) who showed nonuse on the AAUT were identified as at risk for nonuse by the VR-NonUse tool, and 29 out of 33 participants (87.9%) who did not show nonuse on the AAUT were classified as not at risk for nonuse by the VR-NonUse tool. These findings suggest a preliminary threshold of approximately 31% or fewer paretic arm choice during the Complex Grasp condition for identifying individuals at risk for arm nonuse. To further contextualize this threshold in relation to motor impairment, Figure 4 illustrates spontaneous paretic arm choice in the Complex Grasp condition as a function of UEFM scores. Notably, several individuals with mild motor impairment (UEFM>48), based on previously established classification,^46^ demonstrated low spontaneous paretic arm choice despite relatively preserved motor capacity. This observation highlights that low spontaneous paretic arm choice can occur even among individuals with mild motor impairment, supporting the potential utility of the VR-NonUse tool for identifying individuals at risk for arm nonuse despite sufficient motor capacity.

**Figure 4.**
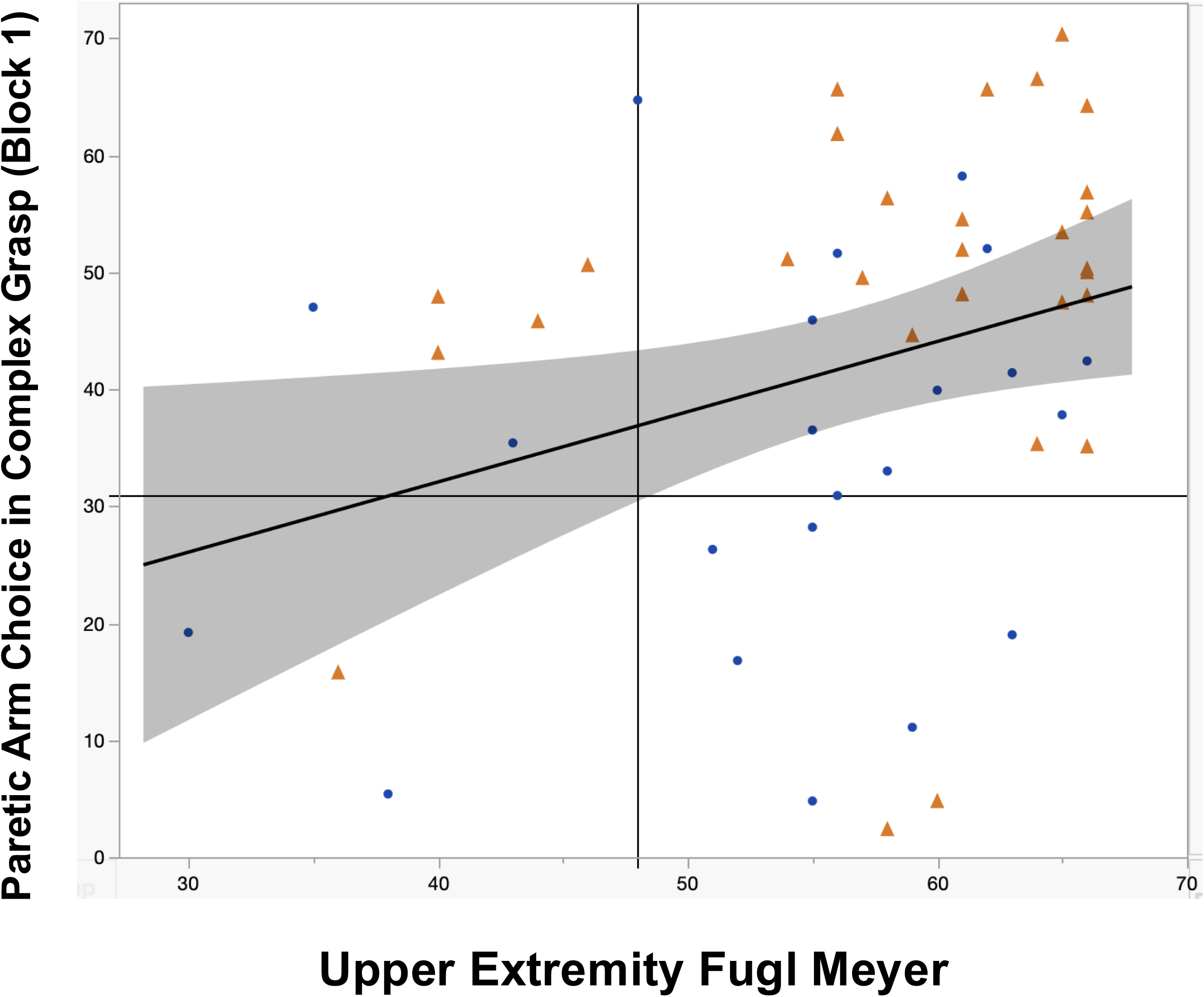
Relationship between Upper Extremity Fugl Meyer (UEFM) and paretic arm choice in Block 1 of the VR-NonUse Complex Grasp condition. Individuals with mild motor impairment (UEFM > 48) who chose their paretic arm on < 31% trials in the Complex Grasp condition (lower right quadrant) may be at risk of nonuse. Participants with RCVA are shown in blue circles; participants with LCVA are shown in orange triangles.

### Post hoc Sensitivity Analyses

To assess sample size sensitivity for the correlation-based validity analyses, we estimated the minimum detectable correlation between spontaneous arm choice on the VR-NonUse tool and established measures of arm nonuse (AAUT Nonuse Index and Actigraph Use Ratio) and motor impairment. With 51 participants with stroke, a two-sided α=.05, and 80% power, the approximate detectable correlation was |*r*|=.38. In the simulation-based sensitivity analysis, power to detect an effect of the magnitude observed for the cognitive-demand comparison (Basic Grasp vs. Complex Grasp) was 96.8% at *N*=74, respectively. For an effect of the magnitude observed for the motor-demand comparison (Complex Grasp vs. Complex Point), estimated power was 30.4% at *N*=74, respectively. Thus, the obtained sample provided high sensitivity to an effect of the magnitude observed for the cognitive-demand comparison but substantially lower sensitivity to the smaller effect observed for the motor-demand comparison.

## Discussion

Our findings indicate that the research version of the VR-NonUse tool has high usability and tolerability, high internal consistency reliability, and moderate concurrent, discriminant, and construct validity for assessing arm nonuse. Towards the goal of item reduction for development of a clinical version of the tool, we demonstrated that the Complex Grasp condition (grasping a target presented among semantically related distractors) is associated with relatively high levels of paretic arm nonuse, particularly in individuals with RCVA. A single block of this condition was sufficient to capture paretic arm nonuse while minimizing assessment burden. We also established a preliminary threshold for paretic arm choice in the Complex Grasp condition of VR-NonUse tool that may be used to identify individuals who are at risk of paretic arm nonuse.

The observed relationships between paretic arm choice on the VR-NonUse tool and established measures of arm nonuse and real-world arm use support its concurrent validity. Less frequent selection of the paretic arm during the VR-NonUse assessment was associated with higher AAUT Nonuse Index and lower accelerometry Use Ratios. These findings suggest that the VR-NonUse tool captures patterns of nonuse consistent with those captured in both controlled (AAUT) and naturalistic (accelerometry) contexts. In addition, the VR-NonUse tool demonstrated statistically stronger relationships with measures of arm nonuse than with motor impairment (UEFM), supporting its discriminant validity. Consequently, the VR-NonUse tool complements conventional impairment-based assessments by identifying individuals whose paretic arm use remains disproportionately reduced relative to their motor function, thereby potentially informing targeted rehabilitation approaches.

The present findings replicate our previous work showing that cognitive demand exacerbates paretic arm nonuse^15^ and provide evidence for the construct validity of the VR-NonUse tool. The increased visual and semantic similarity of objects in the Complex Grasp condition likely increased the cognitive resources required for target detection and selection. Because selecting the paretic arm itself also requires cognitive effort in individuals with stroke,^12,15^ one possible interpretation is that the additional visual and semantic competition in the Complex Grasp condition increased demands on cognitive resources available for paretic arm selection, resulting in greater paretic arm nonuse. This interpretation is consistent with the well-established theory that attention is a limited-capacity resource, whereby increased attentional demands in one task reduce the resources available for other processes.^47^ It is also consistent with evidence that cognitive load impairs motor performance in both neurologically healthy individuals and those with stroke,^48,49^ with disproportionately greater effects after stroke.^50^ These data underscore the importance of incorporating cognitive demand into arm nonuse assessments. A clinical version of the VR-NonUse tool (see below) may help identify individuals who are particularly susceptible to arm nonuse in everyday environments characterized by high attentional demands. Such individuals may benefit from interventions that promote arm use under cognitive load, such as practicing paretic arm use while simultaneously performing cognitive tasks.^51^ Future studies could also examine whether interventions that modify attentional/arousal state, such as psychostimulant medications, influence limb selection and reduce arm nonuse.^52^

In contrast, manipulating motor demand through the pointing-versus-grasping conditions did not influence arm nonuse. However, this finding should be interpreted cautiously, as the simulation-based sensitivity analysis indicated limited power to detect an effect of the magnitude observed for this comparison. Thus, the absence of a statistically significant difference does not preclude smaller, potentially meaningful effects of motor demand on spontaneous paretic arm choice. Additionally, compared with the Complex Point condition, the Complex Grasp condition, showed numerically stronger associations with other measures of arm nonuse. This numerical pattern is consistent with the possibility that grasping more closely reflects the demands of everyday upper-extremity activities than does pointing, perhaps because it requires greater motor planning, hand shaping, and functional coordination.^53–56^ Further investigation with a larger sample will be helpful in determining the influence of motor demand on arm choice behavior.

Across all conditions, individuals with stroke chose their paretic arm less frequently than controls selected their nondominant arm. Consistent with prior work,^13,30^ this difference was particularly pronounced among individuals with RCVA, who selected their paretic arm less frequently than those with LCVA. This finding is consistent with evidence suggesting that right hemisphere lesions disproportionately disrupt spatial attention, body awareness, motor intention, and action monitoring, processes that likely contribute to reduced spontaneous paretic arm choice.^57–59^ The contribution of such attentional and somatosensory factors to arm nonuse is of great interest and is being investigated in a companion study. In addition, because the non-paretic limb is typically the dominant arm following RCVA, greater reliance on that limb may further reduce paretic arm choice. However, although the left arm was the nondominant arm in our right-handed controls and the paretic arm in individuals with RCVA, the RCVA group used the left arm substantially less than controls, indicating that the observed greater arm nonuse after RCVA is unlikely to be attributable solely to nondominance.

All participants completed the assessment without difficulty. We observed no significant differences in arm choice between the first and second blocks of any condition, suggesting the potential for a shorter assessment. Given its sensitivity to arm nonuse, a single block of the Complex Grasp condition (approximately 12–15 minutes) represents a promising approach for developing an abbreviated version of the VR-NonUse tool that captures relevant patterns of arm nonuse while reducing administration burden. The need for such assessments has been noted in the literature.^23^ A shorter protocol may improve feasibility in both research and clinical settings, where time constraints and fatigue effects frequently limit implementation of detailed behavioral assessments. Evaluation of the validity, reliability, and clinical utility of this abbreviated protocol will be required prior to clinical implementation. These considerations will inform the next phase of tool development (Phase 3).^24^

We also conducted an exploratory ROC analysis to establish a preliminary threshold for identifying individuals at risk of arm nonuse. Although there is no established “gold standard” for determining nonuse, we used the published AAUT Nonuse Index as the reference measure, based on a threshold employed in previous work.⁹ The analysis identified a threshold of approximately 31% spontaneous paretic arm choice during the Complex Grasp block that maximized the balance between sensitivity and specificity in distinguishing individuals with and without AAUT-defined nonuse. The VR-NonUse tool demonstrated modest discriminative ability relative to the AAUT, with relatively high specificity but limited sensitivity. The high specificity indicates that the threshold correctly classified most individuals without AAUT-defined nonuse, whereas the limited sensitivity suggests that a substantial proportion of individuals with AAUT-defined nonuse were not identified by the VR-NonUse tool. Given its modest discriminative ability and the absence of an established gold-standard measure of nonuse, the proposed threshold should be considered preliminary and exploratory. Nevertheless, examining spontaneous paretic arm choice using the VR-NonUse tool alongside a measure of motor impairment may provide a clinically relevant approach to characterizing discrepancies between motor capacity and actual arm use. With validation in larger, independent samples, this approach may inform the development of standardized criteria for identifying individuals at risk of arm nonuse.

Several limitations should be considered. First, the study was conducted as part of a larger research project and was not prospectively designed, analyzed, or interpreted according to Consensus-based Standards for the selection of health Measurement Instruments (COSMIN) recommendations.^60^ Second, participants were required to demonstrate a minimum level of upper-extremity motor function, and there were relatively few subjects in the range of moderate impairment. Although motor capacity was analyzed as a continuous variable to account for the full range of observed impairment, the relatively small number of participants with moderate impairments limit the generalizability of the findings to this population. Third, given the sample size, concurrent validity analyses were conducted across the full stroke sample rather than separately for the RCVA and LCVA groups. Future studies with larger samples should examine whether the associations between the VR-NonUse tool and established nonuse and real-world arm use measures differ by lesion hemisphere. Fourth, the proposed cutoff values are exploratory and require validation in a larger, independent cohort before clinical implementation. Fifth, although VR provides experimental control and standardization, it cannot fully capture the complexity of real-world contexts. Future studies should therefore evaluate how well the VR-NonUse tool predicts naturalistic arm use across a broader range of everyday activities and environments, particularly those with high attentional demands. Finally, test-retest reliability was not established and should be examined to support the tool’s use for longitudinal monitoring of spontaneous arm choice and treatment response.

## Conclusion

The VR-NonUse Tool is a valid, reliable, feasible, and potentially clinically useful assessment of arm nonuse. It may identify individuals at risk for arm nonuse who would otherwise go undetected by conventional impairment- or capacity-based assessments. Beyond its clinical utility, the tool also provides a platform for investigating the mechanisms underlying arm nonuse. By systematically manipulating task demands while capturing detailed behavioral and kinematic data, it enables examination of how cognitive, attentional, and motor processes interact to influence arm use after stroke. As such, the VR-NonUse Tool may advance both the identification of arm nonuse and the mechanistic understanding of post-stroke arm nonuse.

## Acknowledgements

We would like to thank Carolee Winstein and Robert Sainburg for their insightful suggestions during the design of the revised VR-NonUse tool and Cory Potts for his input on the analysis of the revised tool. We are grateful to all study participants for their time and contribution to this research.

Author contributions:

## Ethical considerations

This study was approved by the Institutional Review Board of Thomas Jefferson University.

## Consent to participate

Participants provided written informed consent in accordance with the Institutional Review Board of Thomas Jefferson University.

## Consent for publication

Not applicable.

## Data availability

The parent NIH-funded study is ongoing. Upon completion of the study, de-identified data and supporting documentation will be deposited in the NeuroLex/Neuroscience Information Framework (NIF) repository and made publicly available to the research community.

