## Supplemental Table 1 for "A Valid and Reliable Virtual Reality Tool for Assessment of Arm Nonuse Post-Stroke"

**Table 1:** Discriminant Validity: Associations between paretic arm choice on the VR-NonUse Tool conditions with AAUT NU Index, Actigraph Use Ratio and UEFM.

| **Conditions** | **AAUT NU Index** | | | **Actigraph Use Ratio** | | | **UEFM** | | |
| --- | --- | --- | --- | --- | --- | --- | --- | --- | --- |
|  | **ρ** | **p** | **q** | **ρ** | **p** | **q** | **ρ** | **p** | **q** |
| Basic Grasp | -0.40 | 0.003 | 0.004 | 0.53 | <0.0001 | <0.0001 | 0.35 | 0.009 | 0.01 |
| Complex Grasp | -0.46 | 0.0006 | 0.001 | 0.52 | <0.0001 | <0.0001 | 0.34 | 0.01 | 0.01 |
| Complex Point | -0.20 | 0.15 | 0.15 | 0.32 | 0.02 | 0.02 | 0.23 | 0.09 | 0.09 |

AAUT NU Index: Actual Amount of Use Test Nonuse Index; UEFM: Upper Extremity Fugl Meyer, ρ: Spearman rank correlation coefficient; p: uncorrected significance value, q: adjusted p-value controlling the false discovery rate using the Benjamini-Hochberg procedure.

**Figure 1.** Proportion of paretic arm use in Blocks 1 and 2 across the three VR-NonUse conditions.
